# Effectiveness of mRNA COVID-19 monovalent and bivalent vaccine booster doses against Omicron severe outcomes among adults aged ≥50 years in Ontario, Canada

**DOI:** 10.1101/2023.04.11.23288403

**Authors:** Ramandip Grewal, Sarah A Buchan, Lena Nguyen, Sharifa Nasreen, Peter C. Austin, Kevin A. Brown, Jonathan Gubbay, Nelson Lee, Kevin L Schwartz, Mina Tadrous, Kumanan Wilson, Sarah E Wilson, Jeffrey C Kwong, the Canadian Immunization Research Network (CIRN) Provincial Collaborative Network investigators

## Abstract

**Objective:** We estimated the effectiveness of booster doses of monovalent and bivalent mRNA COVID-19 vaccines against Omicron-associated severe outcomes among adults aged ≥50 years in Ontario, Canada.

**Methods:** We used a test-negative design to estimate vaccine effectiveness (VE), with unvaccinated adults as the comparator, against hospitalization or death among SARS-CoV-2-tested adults aged ≥50 years between June 19, 2022 and January 28, 2023 stratified by time since vaccination. We explored VE by vaccine product (Moderna Spikevax^®^ monovalent; Pfizer-BioNTech Comirnaty^®^ monovalent; Moderna Spikevax^®^ BA.1 bivalent; Pfizer-BioNTech Comirnaty^®^ BA.4/BA.5 bivalent).

**Results:** We included 3,755 Omicron cases and 14,338 test-negative controls. For the Moderna and Pfizer-BioNTech monovalent vaccines, VE 7-29 days after vaccination was 85% (95% confidence interval [CI], 72-92%) and 88% (95%CI, 82-92%), respectively, and was 82% (95%CI, 76-87%) and 82% (95%CI, 77-86%) 90-119 days after vaccination. For the Moderna BA.1 bivalent vaccine, VE was 86% (95%CI, 82-90%) 7-29 days after vaccination and was 76% (95%CI, 66-83%) 90-119 days after vaccination. For the Pfizer-BioNTech BA.4/BA.5 bivalent vaccine, VE 7-29 days after vaccination was 83% (95%CI, 77-88%) and was 81% (95%CI 72-87%) 60-89 days after vaccination.

**Conclusions:** Booster doses of monovalent and bivalent mRNA COVID-19 vaccines provided similar, strong initial protection against severe outcomes in community-dwelling adults aged ≥50 years in Ontario. Nonetheless, uncertainty remains around waning protection of these vaccines.

## INTRODUCTION

On September 1, 2022, the Moderna Spikevax^®^ bivalent COVID-19 vaccine targeting the original SARS-CoV-2 virus and the Omicron BA.1 sublineage (Moderna BA.1 bivalent) was approved as a booster dose by Health Canada for adults aged ≥18 years.^1^ The Pfizer-BioNTech Comirnaty^®^ bivalent vaccine targeting the original virus and Omicron BA.4/BA.5 (Pfizer-BioNTech BA.4/BA.5 bivalent) was approved as a booster dose on October 7, 2022 for individuals aged ≥12 years.^2^ Upon their approval in Canada, bivalent COVID-19 vaccines were recommended for booster doses, though monovalent vaccines were still available and accessible.^3^ Canada’s advisory committee on immunization emphasized the importance of receiving a booster dose, regardless of product, to ensure timely protection.^3^ The Moderna BA.1 bivalent vaccine was introduced to Ontario’s COVID-19 vaccination program as a booster dose for adults at highest risk, including adults aged ≥70 years, on September 12, 2022, and was expanded to all adults on September 26, 2022.^4^ The Pfizer-BioNTech BA.4/BA.5 bivalent vaccine was introduced on October 17, 2022.^5^ Among older adults in Ontario, past studies have demonstrated that monovalent mRNA COVID-19 vaccines provide strong protection against Omicron-related severe outcomes with evidence of some waning against protection over time^6,7^ but the effectiveness of bivalent vaccines has not yet been explored. Our objective was to compare the vaccine effectiveness (VE) of booster vaccinations using the Moderna BA.1 and Pfizer-BioNTech BA.4/BA.5 bivalent mRNA COVID-19 vaccines and the original mRNA COVID-19 monovalent vaccines against Omicron-related severe outcomes in Ontario, Canada.

## METHODS

We conducted a test-negative design study among community-dwelling adults aged ≥50 years who had ≥1 real-time polymerase chain reaction (RT-PCR) test for SARS-CoV-2 between June 19, 2022 and January 28, 2023. We used provincial SARS-CoV-2 laboratory testing, COVID-19 surveillance, COVID-19 vaccination, and health administrative datasets, which were linked using unique encoded identifiers and analyzed at ICES (formerly the Institute for Clinical Evaluative Sciences). Projects that use data collected by ICES under section 45 of PHIPA, and use no other data, are exempt from research ethics board review.

Among community-dwelling adults, we excluded immunocompromised adults (n=10,715), adults who received <4 doses (n=40,422) or received a vaccine not authorized by Health Canada (n=13), those who tested positive within 60 days prior to the index test (n=36), and hospitalizations when specimen collection occurred >3 days after admission (n=505) or if flagged as nosocomial (n=388). Third doses, or first booster doses, were excluded due to the significant time gap between third dose introduction (December 2021) and bivalent vaccine introduction (September 2022) in Ontario. Adults presenting for ≥2 monovalent booster doses were considered to be more similar to those presenting for a bivalent booster dose. Sublineage-predominant periods were defined as ≥50% of sequenced samples in the province being of that sublineage. From June 19, 2022 to December 3, 2022, Omicron BA.4/BA.5 was the predominant sublineage and from December 4, 2022 to January 28, 2023, BQ was predominant (Supplementary Appendix, Figure S1).^8^

Compared to unvaccinated subjects, we estimated VE of Moderna monovalent (mRNA-1273 [50 μg ancestral Wuhan-Hu-1]), Pfizer-BioNTech monovalent (BNT162b2 [30 μg ancestral Wuhan-Hu-1]), Moderna BA.1 bivalent (mRNA-1273.214 [25 μg each of ancestral Wuhan-Hu-1 and Omicron BA.1]), and Pfizer-BioNTech BA.4/BA.5 bivalent (15 μg each of ancestral Wuhan-Hu-1 and Omicron BA.4/BA.5) booster doses (dose 4 or greater) against severe outcomes (hospitalization or death due to, or partially due to, COVID-19) stratified by time since vaccination (30-day periods up to 4 months). The Moderna BA.4/BA.5 bivalent vaccine was approved in Canada during the study period but the vaccine was not yet available in Ontario. Cases and controls were sampled by week of test. Once an individual became a case, they could no longer re-enter the study.

We used multivariable logistic regression to compare the odds of vaccination in cases to test-negative controls. Few adults appeared in models more than once (<10%) with minimal impact on confidence intervals, thus clustering methods were not applied. Models were adjusted for sex, age, prior influenza vaccination (proxy for health behaviours), public health unit region, socio-demographic area-level variables (household income quintile, essential worker quintile, persons per dwelling quintile, self-identified visible minority quintile), number of SARS-CoV-2 tests within 3 months prior to December 14, 2020 (proxy for healthcare workers), comorbidities, receipt of home care services, and week of test. VE of the Pfizer-BioNTech BA.4/BA.5 bivalent vaccine could only be estimated up to 89 days post-vaccination due to its more recent availability in Ontario. We calculated VE using the formula 1-adjusted odds ratio. As a secondary analysis, we stratified our study period by predominant sublineage (BA.4/BA.5 versus BQ).

## RESULTS

We included 3,755 Omicron cases and 14,338 test-negative controls (16,247 unique individuals). Cases were older than controls (mean age 80 years versus 72 years) (Table 1). Approximately 57% of cases and 69% of controls had their fourth dose as their most recent dose. Compared to unvaccinated adults, fewer vaccinated adults were male and from an area with the lowest household income quintile (Supplementary Appendix, Table S2). Significantly more vaccinated adults had a recent prior influenza vaccine. From June 19 to September 11, 2022, when only monovalent COVID-19 vaccines were available, 33% and 66% of adults received the Moderna monovalent and Pfizer-BioNTech monovalent vaccine, respectively, as their most recent dose. From September 12, 2022 onward, 16%, 18%, 31%, and 35% received the Moderna monovalent, Pfizer-BioNTech monovalent, Moderna BA.1 bivalent, and Pfizer-BioNTech BA.4/BA.5 bivalent vaccine, respectively, as their most recent dose.

**Table 1.**
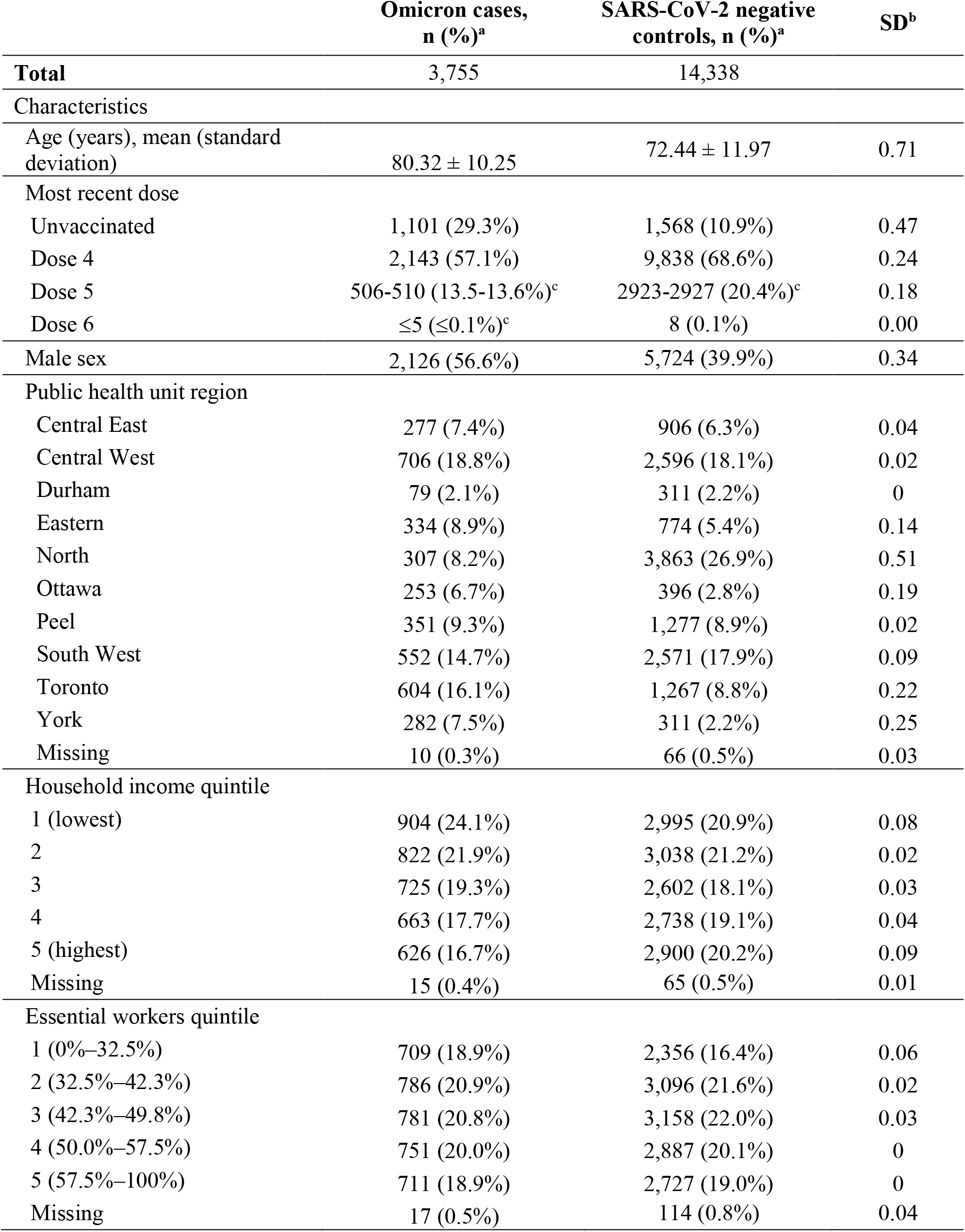
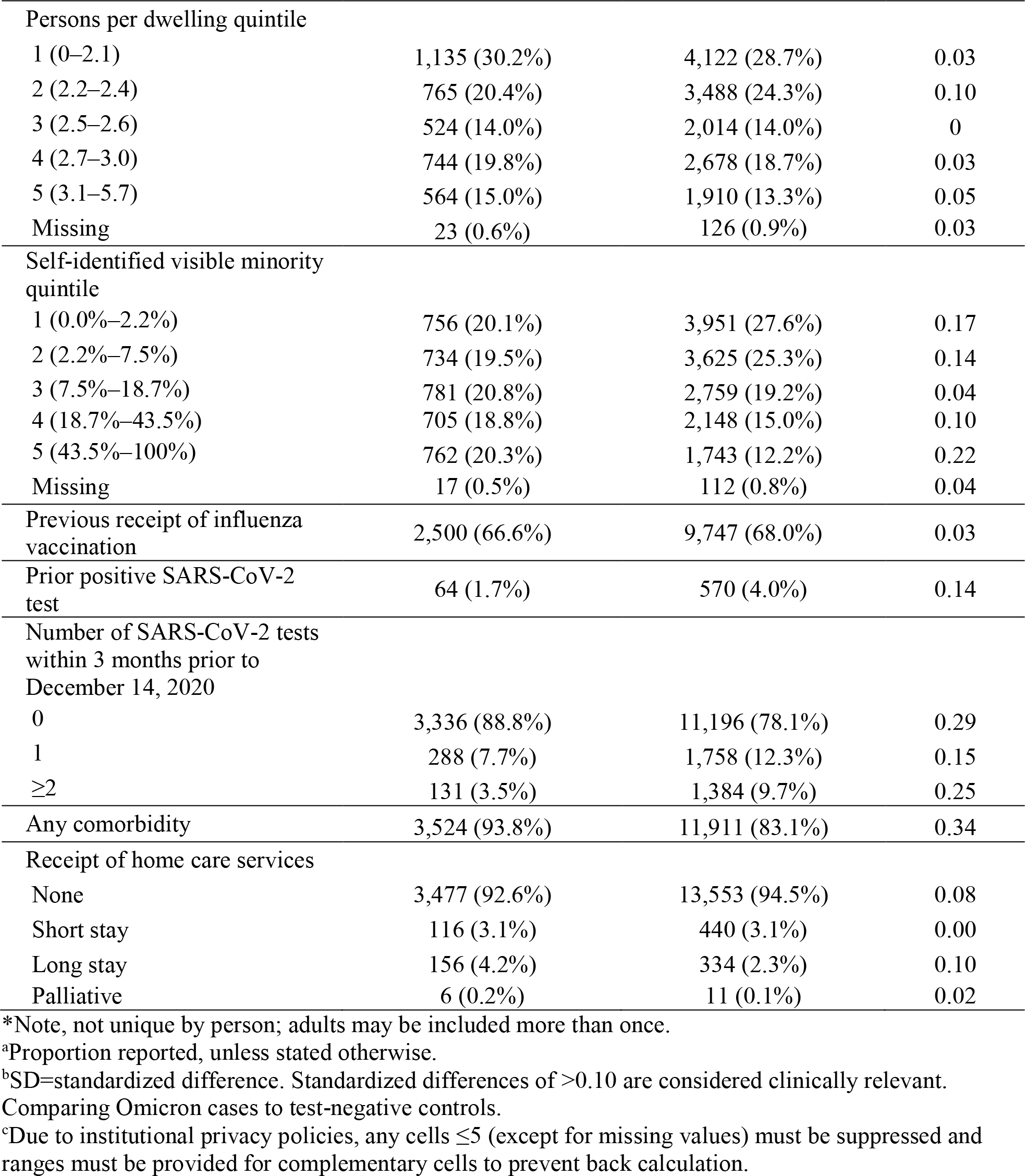
Descriptive characteristics of community-dwelling adults aged ≥50 years tested for SARS-CoV-2 between June 19, 2022 and January 28, 2023 in Ontario, Canada, comparing Omicron-associated severe outcome cases to SARS-CoV-2 negative controls

For the full study period, VE of the Moderna and Pfizer-BioNTech monovalent vaccines 7-29 days after vaccination was 85% (95% confidence interval [CI], 72-92%) and 88% (95%CI 82-92%), respectively (Figure 1; Supplementary Appendix, Table S3). VE at 90-119 days after vaccination was 82% (95%CI, 76-87%) for the Moderna monovalent vaccine and 82% (95%CI, 77-86%) for the Pfizer-BioNTech monovalent vaccine. VE of the Moderna BA.1 bivalent vaccine was 86% (95%CI, 82-90%) 7-29 days after vaccination and 76% (95%CI, 66-83%) 90-119 days after vaccination. VE of the Pfizer-BioNTech BA.4/BA.5 bivalent vaccine was 83% (95%CI, 77-88%) 7-29 days after vaccination and 81% (95%CI, 72-87%) 60-89 days after vaccination.

**Figure 1:**
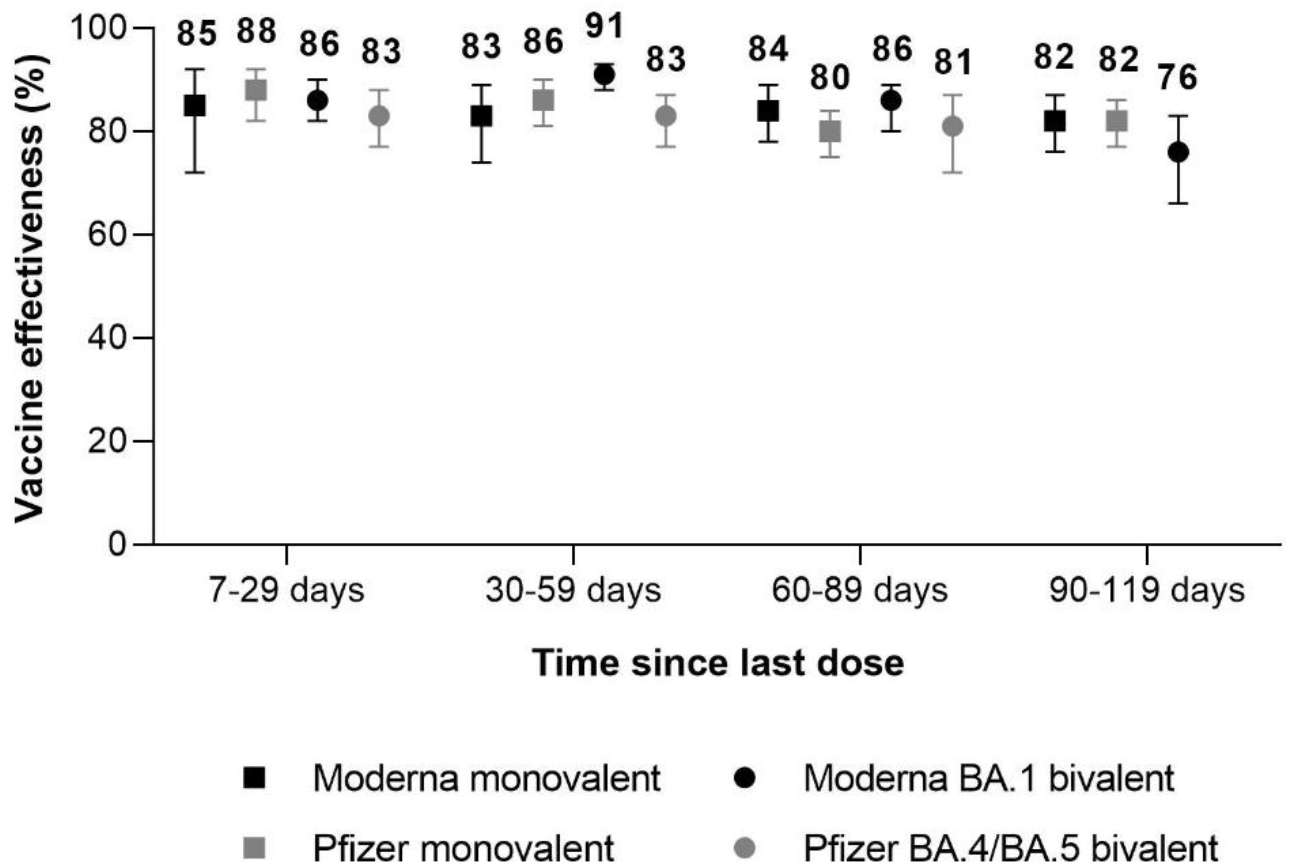
Vaccine effectiveness of monovalent and bivalent mRNA COVID-19 vaccines against Omicron-associated severe outcomes by time since vaccination among community-dwelling adults aged ≥50 years in Ontario, Canada, compared to unvaccinated adults, June 19, 2022 to January 28, 2023 (BA.4/BA.5 and BQ periods combined).

Comparisons of all vaccine products and time intervals since vaccination were not possible when stratified by sublineage period due to different dates of vaccine introduction. For estimates that could be compared, VE was slightly lower during the BQ-predominant period than the BA.4/BA.5-predominant period (Figure 2; Supplementary Appendix, Table S4). For example, VE of the Moderna BA.1 bivalent vaccine and Pfizer-BioNTech BA.4/BA.5 bivalent vaccine was 93% (95%CI, 90-95%) and 87% (95%CI, 71-94%), respectively, 30-59 days after vaccination during the BA.4/BA.5 predominant period, whereas the corresponding estimates during the BQ-predominant period were 82% (95%CI, 71-89%) and 82% (95%CI, 73-88%).

**Figure 2:**
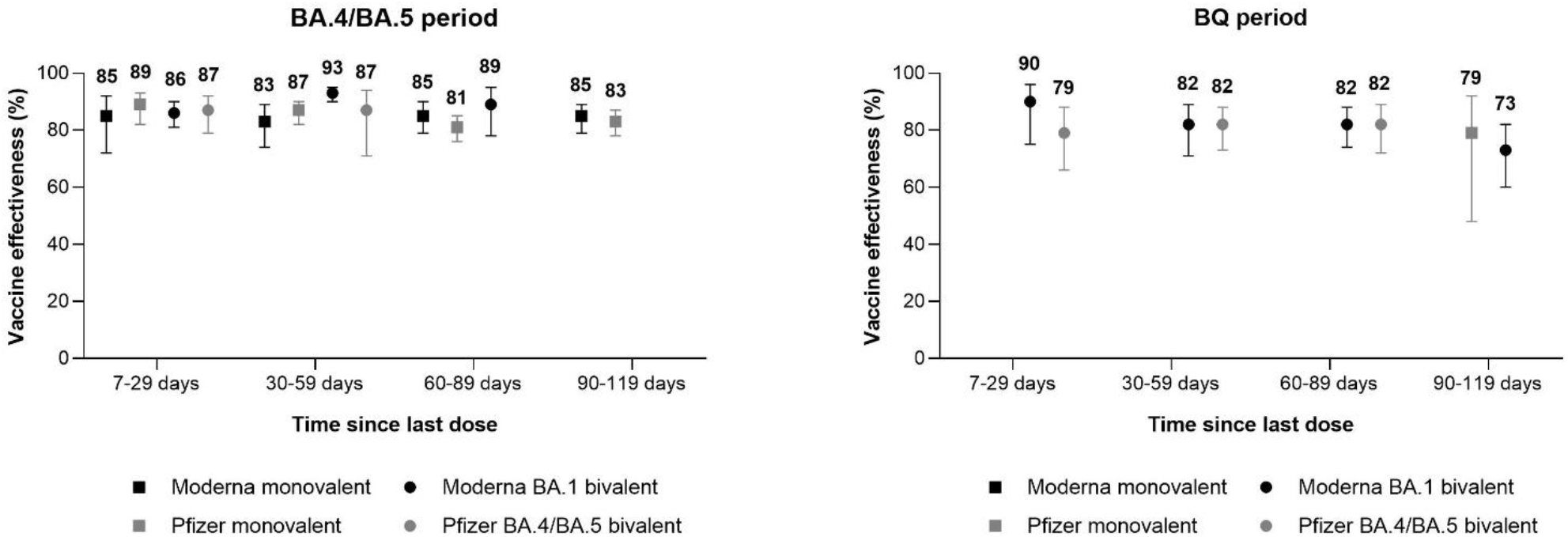
Vaccine effectiveness of monovalent and bivalent mRNA COVID-19 vaccines against Omicron-associated severe outcomes by time since vaccination among community-dwelling adults aged ≥50 years in Ontario, Canada, compared to unvaccinated adults, stratified by sublineage period (BA.4/BA.4 period = June 19 to December 3, 2022; BQ period = December 4, 2022 to January 28, 2023).

## DISCUSSION

Among community-dwelling adults aged ≥50 years in Ontario, monovalent and bivalent mRNA COVID-19 vaccines provided similar, strong initial protection against hospitalization or death. VE of monovalent and bivalent vaccines ranged from 85-88% and 83-86%, respectively, 7-29 days after vaccination. There was only slight waning of protection against severe outcomes across the 4-month period. After 90-119 days, VE of monovalent vaccines decreased to 82% and VE of the Moderna BA.1 bivalent vaccine decreased to 76%. VE appeared slightly lower in the BQ-predominant period compared to the BA.4/BA.5 period where comparisons were possible, though uncertainty remains whether a true difference exists. A potentially lower VE against BQ may contribute to lower VE in latter time periods versus waning of protection alone.

There are few studies comparing the VE of a subsequent dose of a COVID-19 monovalent versus bivalent vaccine across a similar time period. Comparable to our findings, the few existing studies have suggested that the effectiveness of bivalent vaccines is either similar or moderately higher than the monovalent products.^9–12^ Nonetheless, due to differences in circulating sublineages at the time of vaccine introduction and across the study period (e.g., prevalence of a sublineage will differ even across 30-day intervals) as well as differences in rates of previous infection, direct comparisons of effectiveness between monovalent and bivalent vaccines within and across jurisdictions is difficult.

There is limited evidence on the VE of monovalent and bivalent COVID-19 vaccines against BQ-associated outcomes, particularly severe disease. Similar to our results, findings from England suggested that bivalent VE was lower against BQ than against BA.5-related hospitalizations 2 or more weeks after vaccination (∼10 percentage point difference).^13^ The study also expressed uncertainty around whether a difference really exists.

There are limitations to this analysis. Evidence suggests that VE of COVID-19 vaccines differs among those with versus without a prior SARS-CoV-2 infection.^14^ We were unable to account for prior infection as an effect modifier in our analysis since few adults had a prior infection in our data. Due to limited RT-PCR testing availability in Ontario and no access to rapid antigen testing results, the majority of SARS-CoV-2 infections in the province are undocumented. If unvaccinated adults were more likely to have a previous infection than vaccinated adults, VE may be underestimated since unvaccinated adults would have some underlying existing infection-induced immunity. Nonetheless, increasing SARS-CoV-2 seroprevalence in Ontario (76% among adults as of January 15, 2023)^15^ suggests that VE estimates may be more generalizable toward those that have been previously infected. There is also potential for residual confounding since we were limited to the variables available in our datasets. A significant strength of our analysis is that due to the continued availability and uptake of monovalent mRNA COVID-19 vaccines after bivalent vaccines were introduced, we were able to compare the VE of these products as subsequent booster doses across a similar study period.

Monovalent and bivalent mRNA COVID-19 vaccines provide comparable levels of initial protection against Omicron-related severe outcomes among community-dwelling adults aged ≥50 years in Ontario. Older adults continue to experience the highest rates of COVID-19 related severe outcomes and would benefit the most with additional vaccine doses.^16^ Longer follow-up is necessary to determine the long-term protection of bivalent vaccines and the effectiveness against newer Omicron sublineages such as XBB.

## Data Availability

The dataset from this study is held securely in coded form at ICES. While legal data sharing agreements between ICES and data providers (e.g., healthcare organizations and government) prohibit ICES from making the dataset publicly available, access may be granted to those who meet prespecified criteria for confidential access, available at www.ices.on.ca/DAS. The full dataset creation plan and underlying analytic code are available from the authors upon request, understanding that the computer programs may rely upon coding templates or macros that are unique to ICES and are therefore either inaccessible or may require modification.

## Acknowledgements

This work was supported by funding from the Canadian Immunization Research Network (CIRN) through a grant from the Public Health Agency of Canada and the Canadian Institutes of Health Research (CNF 151944), and also by funding from the Public Health Agency of Canada, through the Vaccine Surveillance Working Party and the COVID-19 Immunity Task Force. This study was supported by Public Health Ontario and by ICES, which is funded by an annual grant from the Ontario Ministry of Health (MOH) and Ministry of Long-Term Care (MLTC). This work was also supported by the Ontario Health Data Platform (OHDP), a Province of Ontario initiative to support Ontario’s ongoing response to COVID-19 and its related impacts. Jeffrey C. Kwong is supported by a Clinician-Scientist Award from the University of Toronto Department of Family and Community Medicine. The study sponsors did not participate in the design and conduct of the study; collection, management, analysis and interpretation of the data; preparation, review or approval of the manuscript; or the decision to submit the manuscript for publication.

We would like to acknowledge the Canadian Immunization Research Network (CIRN) Provincial Collaborative Network (PCN) Investigators, Public Health Ontario for access to vaccination data from COVaxON, case-level data from the Public Health Case and Contact Management Solution (CCM) and COVID-19 laboratory data, as well as assistance with data interpretation. We also thank the staff of Ontario’s public health units who are responsible for COVID-19 case and contact management and data collection within CCM. We thank IQVIA Solutions Canada Inc. for use of their Drug Information File. The authors are grateful to the Ontario residents without whom this research would be impossible.

This document used data adapted from the Statistics Canada Postal Code^OM^ Conversion File, which is based on data licensed from Canada Post Corporation, and/or data adapted from the Ontario Ministry of Health Postal Code Conversion File, which contains data copied under license from ^©^Canada Post Corporation and Statistics Canada. Parts of this material are based on data and/or information compiled and provided by: MOH, Ontario Health, the Canadian Institute for Health Information, Statistics Canada, and IQVIA Solutions Canada Inc. The analyses, conclusions, opinions and statements expressed herein are solely those of the authors and do not reflect those of the funding or data sources; no endorsement is intended or should be inferred. Adapted from Statistics Canada, Canadian Census 2016. This does not constitute an endorsement by Statistics Canada of this product.

